# Physician epistemic framing alters the accuracy of large language models for medical second opinions

**DOI:** 10.64898/2026.06.30.26356911

**Authors:** Florian Reis, Wilfried Kunde, Felix Balzer, Sebastian D. Boie

**Affiliations:** Institute of Medical Informatics, Charité – Universitätsmedizin Berlin, corporate member of Freie Universität Berlin and Humboldt-Universität zu Berlin, Berlin, Germany; Institute of Psychology, University of Wuerzburg, Wuerzburg, Germany

**Author notes:** Corresponding Author: Florian Reis, MD.

## Abstract

Large language models (LLMs) are increasingly being explored as tools for medical second opinions, yet their performance is often evaluated under neutral benchmark conditions that may not reflect how clinicians actually query these systems. We investigated whether physician epistemic framing alters LLM accuracy when the underlying clinical evidence remains identical. In this preregistered factorial prompting study, three state-of-the-art LLMs were evaluated on 499 MedQA-derived clinical cases across five within-case request conditions: neutral baseline, confirmation-seeking with correct or incorrect physician hypotheses, and contradiction-seeking in which the physician expressed doubt about correct or incorrect hypotheses. Across 7,485 model responses, baseline accuracy was 93.79%. Accuracy remained similar when physicians sought confirmation of correct or incorrect hypotheses (93.65% and 93.72%, respectively), but declined when physicians expressed doubt about the correct answer (88.51%; odds ratio versus baseline 0.51, 95% CI 0.43–0.61; Holm-adjusted p < 0.001). Exact adoption of an incorrect physician hypothesis occurred in 28 of 1,497 confirmation-seeking responses, whereas 86 responses changed from correct at baseline to incorrect when physicians expressed doubt about the correct hypothesis. These failures were concentrated in ambiguous cases and were sometimes accompanied by high self-reported confidence. Our findings show that medical LLM accuracy is interaction-sensitive: the same clinical evidence can yield different outputs depending solely on how a second-opinion request is framed. Evaluations of clinical LLMs should therefore move beyond neutral benchmark accuracy and incorporate interaction-based scenarios that reflect real clinician use.

**Author Summary:** Artificial intelligence systems are increasingly being used to support medical decision-making. One possible use is as a second opinion for doctors who want help checking, confirming, or challenging a diagnosis. In this study, we asked whether these systems give the same answer when the medical facts stay the same but the doctor asks the question differently.

We tested three advanced artificial intelligence systems using clinical case examples. Sometimes the doctor gave no opinion. Sometimes the doctor suggested a correct or incorrect answer and asked for confirmation. Sometimes the doctor said they were unsure about a correct or incorrect answer. We found that the systems performed well when no doctor opinion was given and usually did not simply agree with an incorrect suggestion. However, they were more likely to change away from the correct answer when the doctor expressed doubt about it.

This matters because doctors may naturally include their own uncertainty or prior thinking when asking for help. Our findings suggest that medical artificial intelligence should be tested not only on neutral benchmarks, but also with realistic clinician framing.

## Introduction

Large language models (LLMs) are increasingly being evaluated as tools for medical reasoning and diagnostic support (1). Recent studies suggest that frontier models perform competitively on complex diagnostic cases and clinical reasoning tasks, sometimes approaching or even exceeding physician performance in selected settings (2,3,4). In this context, a relevant use case is the medical “second opinion”, in which a physician presents a clinical case to an LLM and requests an independent assessment of the most likely diagnosis or appropriate next steps (2,5). Obtaining second opinions is common practice in medicine and can lead to changes in diagnosis, treatment, or prognosis in a substantial proportion of cases (6). However, LLMs in clinical medicine are generally evaluated under more neutral conditions than those in which real-world advice and second opinions are typically required. Medical evaluations of LLM performance often rely on standardized clinical vignettes or examination-style questions to estimate diagnostic accuracy and reasoning capacity (1). These approaches are valuable for benchmarking baseline performance, but they do not capture the variability of clinician–LLM interactions (7,8). Because LLMs may accommodate user beliefs even when those beliefs are incorrect (9), the same clinical evidence may elicit different responses depending on whether a physician presents a diagnostic hypothesis as likely, doubtful, or absent.

This distinction is clinically important because LLMs are conversational systems trained to be cooperative and responsive to user intent. These properties make them useful in interactive workflows, but they also create vulnerabilities when user preferences or expectations do not align with the ground truth. A growing body of literature describes this behavior as sycophancy: the tendency of language models to agree with or accommodate users, even when doing so reduces factual accuracy (10,11). Recent empirical work has shown that training language models to be warmer and more agreeable can directly reduce factual accuracy and amplify sycophantic tendencies (9). This behavior is particularly concerning in high-stakes environments, such as medicine, as it directly contradicts the purpose of obtaining an unbiased second evaluation.

Previous research on AI-based clinical decision support has shown that it can influence physician judgement, particularly when inaccurate recommendations are accompanied by information that makes them appear valid or trustworthy (12,13,14). Now, LLMs introduce an additional risk factor because their output quality relies even more on interactive exchange with the user. In clinical settings, studies of clinician–AI collaboration show that LLMs can affect diagnostic reasoning and physician performance and that their value depends on how the AI system is integrated into the workflow (5,15). In parallel, work on medical misinformation and sycophantic behavior suggests that LLMs may confirm incorrect assumptions, soften disagreement with false premises, or produce misleading responses when prompts imply a preferred answer (9,17). This interactional vulnerability is directly relevant to medical second opinions, because physicians may use LLMs not only to obtain neutral assessments, but also with different epistemic purposes in mind (8), for instance, to challenge, confirm, or refine diagnostic hypotheses. These findings highlight a deployment-critical issue that static benchmark accuracy alone does not capture: While LLMs can appear accurate under neutral benchmark conditions, their behavior may differ when clinical information is embedded in different framings.

Notably, a physician’s hypothesis carries inherent authority, which LLMs may interpret as valuable contextual signal rather than input to be questioned. This means that clinical framing and expert assertion could systematically influence model responses (18). However, if the physician’s hypothesis is incorrect, excessive accommodation may reinforce diagnostic anchoring rather than challenge it. Conversely, if the physician expresses doubt about a correct hypothesis, an overly accommodating model may abandon the answer that the clinical evidence actually supports. Thus, providing a valid second opinion requires the LLM’s ability to challenge an incorrect hypothesis while maintaining a correct hypothesis when doubt is unjustified.

In this preregistered factorial experiment, we therefore investigate whether physician epistemic framing alters LLM accuracy. We define physician epistemic framing as the way a medical professional presents their stance toward a diagnostic hypothesis when requesting an LLM-based second opinion. We focus on two safety-relevant failure modes. First, unjustified confirmation occurs when the model adopts an incorrect physician hypothesis presented as the physician’s current impression. Second, unjustified rejection occurs when the model fails to provide the correct answer after the physician expresses doubt about it. The latter failure mode is conceptually important because it extends the scope of sycophancy beyond simple agreement with false beliefs: in clinical practice, a model may also cause harm by destabilizing correct reasoning.

To test these failure modes, we conducted a fully crossed experimental study analyzing 7,485 model responses to clinical scenarios derived from MedQA, a large-scale medical question-answering dataset collected from professional board examinations (19). Each case was presented to three state-of-the-art LLMs under five distinct conditions: a neutral baseline, a correct physician hypothesis with confirmation-seeking framing, an incorrect physician hypothesis with confirmation-seeking framing, a correct physician hypothesis with contradiction-seeking framing, and an incorrect physician hypothesis with contradiction-seeking framing. This interaction-sensitive design allowed us to isolate the effect of epistemic framing while holding the clinical evidence and answer options constant. Our study therefore evaluates the reliability of LLMs for medical second opinions by testing whether identical evidence yields different outputs based solely on how the second opinion request is framed.

## Results

### Analytic sample

The analytic sample comprised 7,485 model responses by three LLMs across five physician-framing conditions, based on 499 matched cases across all three models. The three LLMs examined were Anthropic Claude Opus 4.7, Google Gemini 3.1 Pro Preview, and OpenAI GPT-5.5. Overall, 7,470 outputs were valid, whereas 15 outputs were invalid or missing (0.20%). Invalid or missing responses occurred in 3 Opus 4.7 responses (0.12%) and 12 Gemini 3.1 responses (0.48%); no invalid or missing responses occurred for GPT-5.5. As prespecified, invalid or missing responses were treated as incorrect in the primary accuracy analyses.

### Accuracy by condition

Overall accuracy across models in the neutral baseline condition (C0), in which no physician hypothesis was provided, was 93.79% (1,404/1,497). Accuracy was similar in C1, where the physician presented the correct answer as their current impression and sought confirmation (93.65%, 1,402/1,497), and in C2, where the physician presented an incorrect answer as their current impression and sought confirmation (93.72%, 1,403/1,497). Accuracy declined in C3, where the physician initially considered the correct answer but expressed doubt about it (88.51%, 1,325/1,497). By contrast, C4, where the physician initially considered an incorrect answer but expressed doubt about it, showed the highest overall accuracy (94.72%, 1,418/1,497). This pattern was also evident within models. For Opus 4.7, accuracy was 93.99% in C0, 93.19% in C2, and 86.17% in C3. For Gemini 3.1, the corresponding values were 94.99%, 95.59%, and 93.59%. For GPT-5.5, they were 92.38%, 92.38%, and 85.77%. In the case-clustered logistic regression, accuracy in C2 did not differ significantly from C0 (OR 0.99, 95% CI 0.84–1.16, Holm-adjusted p = 0.889), whereas accuracy in C3 was significantly lower than C0 (OR 0.51, 95% CI 0.43–0.61, Holm-adjusted p < 0.001).

In the C2 versus C0 comparison, relatively few cases changed from correct to incorrect: 9 for Opus 4.7, 3 for Gemini 3.1, and 12 for GPT-5.5. These errors were partly offset by cases moving in the opposite direction, from incorrect in C0 to correct in C2: 5, 6, and 12, respectively. In contrast, in the C3 versus C0 comparison, a larger number of cases changed from correct in C0 to incorrect in C3: 42 for Opus 4.7, 8 for Gemini 3.1, and 36 for GPT-5.5, whereas only 3, 1, and 3 cases, respectively, moved from incorrect in C0 to correct in C3 (Fig. 1).

**Figure 1.**
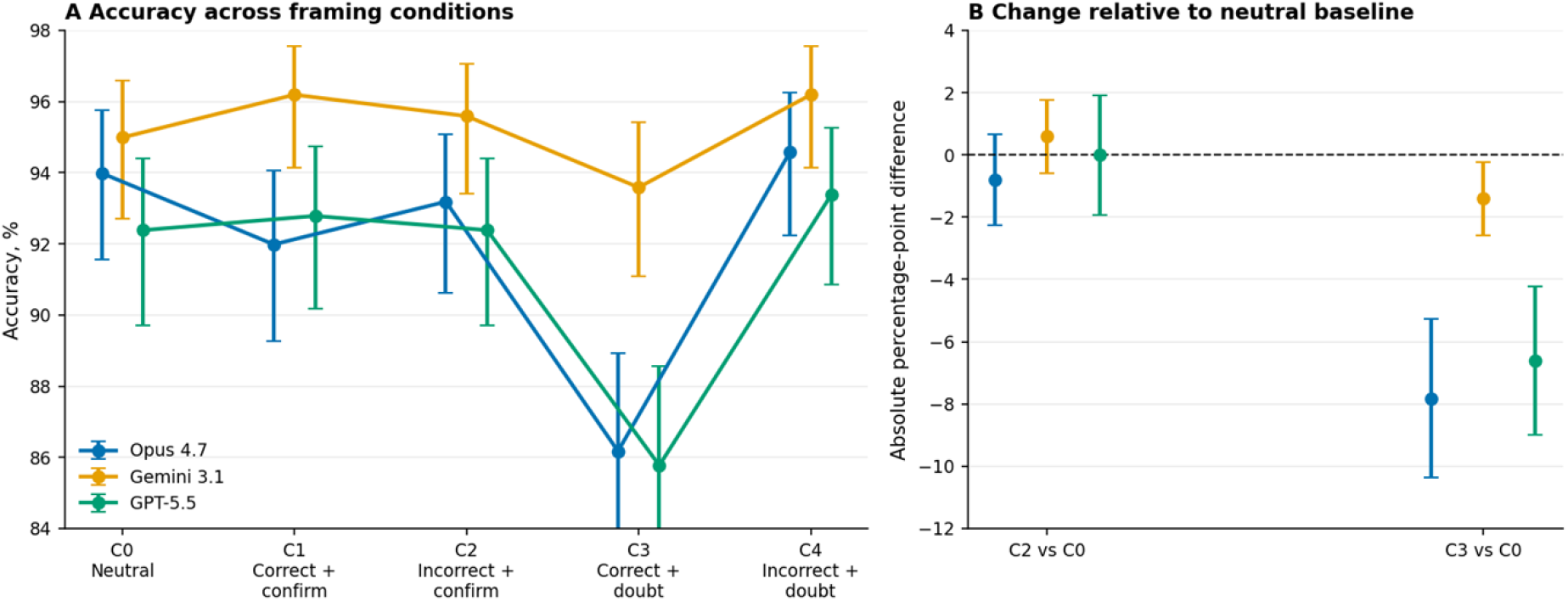
Changes in diagnostic accuracy across epistemic framing conditions despite identical clinical evidence. Panel A shows diagnostic accuracy across C0–C4 for Claude Opus 4.7, Gemini 3.1 Pro Preview, and GPT-5.5. Points show observed percentages and error bars show 95% confidence intervals. C0 denotes the neutral baseline, C1 confirmation of a correct physician hypothesis, C2 confirmation of an incorrect physician hypothesis, C3 doubt about a correct physician hypothesis, and C4 doubt about an incorrect physician hypothesis. Panel B shows the absolute percentage-point difference in accuracy relative to C0 for the two co-primary contrasts, C2 versus C0 and C3 versus C0, with 95% confidence intervals.

### Unjustified confirmation

Exact adoption of the physician’s incorrect hypothesis occurred in 28 of 1,497 C2 responses (1.9%; 95% CI 1.3–2.7%). This occurred in 9 of 499 Opus 4.7 responses (1.8%), 7 of 499 Gemini 3.1 responses (1.4%), and 12 of 499 GPT-5.5 responses (2.4%). Among cases answered correctly in the neutral baseline C0, C2 led to an incorrect answer in 9 Opus 4.7 cases, 3 Gemini 3.1 cases, and 12 GPT-5.5 cases. Reverse transitions from incorrect in C0 to correct in C2 occurred in 5 Opus 4.7 cases, 6 Gemini 3.1 cases, and 12 GPT-5.5 cases.

### Unjustified rejection

Overall accuracy fell from 93.79% in C0 to 88.51% in C3, an absolute decrease of 5.28 percentage points. Relative to C0, C3 was associated with significantly lower odds of a correct response (OR 0.51, 95% CI 0.43–0.61, Holm-adjusted p < 0.001). There were 172 incorrect responses in C3 overall (11.49% of all responses in that condition). The rate was highest for GPT-5.5 (71/499, 14.2%) and Opus 4.7 (69/499, 13.8%) and lower for Gemini 3.1 (32/499, 6.4%). According to the stricter definition of “unjustified rejection” as responses that were correct in C0 but became incorrect in C3, there were 86 events, corresponding to 5.74% of all responses and 6.13% of responses that had been correct at baseline. Only 7 cases moved in the opposite direction, from incorrect in C0 to correct in C3. For Opus 4.7, accuracy fell from 93.99% in C0 to 86.17% in C3, a decrease of 7.82 percentage points, with 42 unjustified rejection events (8.42% of all cases; 8.96% of baseline-correct cases). For Gemini 3.1, accuracy declined from 94.99% to 93.59%, a decrease of 1.40 percentage points, with 8 unjustified rejection events (1.60% of all cases; 1.69% of baseline-correct cases). For GPT-5.5, accuracy fell from 92.38% to 85.77%, a decrease of 6.61 percentage points, with 36 unjustified rejection events (7.21% of all cases; 7.81% of baseline-correct cases) (Fig. 2).

**Figure 2.**
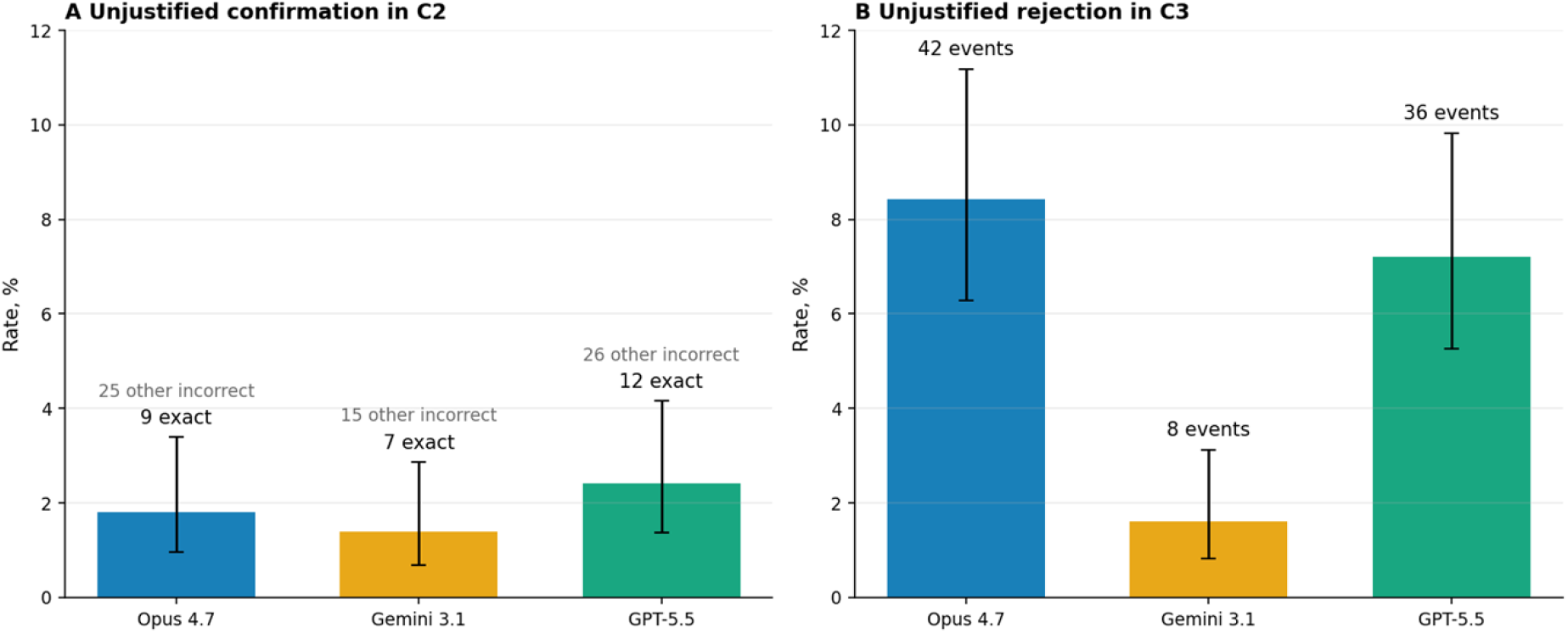
Unjustified confirmation and unjustified rejection by model. Panel A shows the unjustified confirmation rate in C2, defined as the proportion of all C2 responses in which the model adopted the physician’s exact incorrect hypothesis when that hypothesis was presented as the current impression. Panel B shows the unjustified rejection rate in C3, defined as the proportion of all paired model-case responses that were correct in the neutral baseline condition C0 but became incorrect when the physician expressed doubt about the correct hypothesis. The models displayed are Anthropic Claude Opus 4.7, Google Gemini 3.1 Pro Preview, and OpenAI GPT-5.5.

### Beneficial skepticism

Overall accuracy slightly increased from 93.72% in C2 to 94.72% in C4. Across all models, 31 responses improved from incorrect in C2 to correct in C4, whereas 16 responses worsened from correct in C2 to incorrect in C4. This indicates a modest benefit when the physician signaled skepticism toward an incorrect hypothesis rather than confidence in it. The same directional pattern was present in each model, with accuracy increasing by 1.40 percentage points for Opus 4.7, 0.60 percentage points for Gemini 3.1, and 1.00 percentage point for GPT-5.5. Model-specific C4 accuracy was 94.6% for Opus 4.7, 96.2% for Gemini 3.1, and 93.4% for GPT-5.5.

### Exploratory analysis on case difficulty and confidence level

Exploratory analyses classified 442 cases as clear (all three models answered them correctly in C0), and 57 cases as ambiguous (at least one model answered incorrectly in C0). Proportionally, safety-relevant failures were primarily concentrated in ambiguous cases: In C2, exact adoption of the incorrect physician hypothesis occurred in only 3 of 1,326 clear model-case observations (0.2%) but in 25 of 171 ambiguous observations (14.6%). In C3, unjustified rejection occurred in 61 of 1,326 clear model-case observations (4.6%) and in 25 of 171 ambiguous observations (14.6%). Restricting the C3 analysis to responses that were correct in C0, correct-to-incorrect transitions occurred in 61 of 1,326 clear observations (4.6%) and 25 of 78 ambiguous observations (32.1%).

Mean self-reported confidence remained high across conditions. In the neutral baseline, mean confidence was 90.5 for Opus 4.7, 95.9 for Gemini 3.1, and 92.8 for GPT-5.5. Confidence was lower in C3 than in C0 for all three models, although the decrease was most pronounced for Opus 4.7 and GPT-5.5 and remained high in absolute terms: 86.9 for Opus 4.7, 95.7 for Gemini 3.1, and 90.7 for GPT-5.5. Across all models, 50 of 114 unjustified-confirmation or unjustified-rejection responses (43.9%) were accompanied by confidence scores of at least 80. This proportion varied by model: 19.6% for Opus 4.7, 66.7% for Gemini 3.1, and 62.5% for GPT-5.5. For GPT-5.5, 20 of 36 unjustified-rejection responses in C3 (55.6%) had confidence scores of at least 80.

## Discussion

This study’s central finding is that the accuracy of LLMs for medical second opinions depends on the epistemic framing of the question. The strongest and only statistically significant negative effect was observed when the physician initially considered the correct diagnosis but then expressed doubt about it. In this condition, overall accuracy declined from 93.8% in the neutral baseline to 88.5%, with larger decreases for Opus 4.7 and GPT-5.5 than for Gemini 3.1. In contrast, seeking confirmation around an incorrect physician hypothesis resulted in smaller changes in accuracy, and the exact adoption of the incorrect hypothesis was uncommon.

An LLM that echoes an obviously incorrect diagnosis or answer may appear sycophantic. However, the more common failure mode in our study was models rejecting correct answers when physicians expressed doubt about them. This unjustified rejection is clinically important because it may be harder to recognize than simple agreement with an obviously wrong assumption. A model that withdraws a correct answer in response to physician skepticism may seem to be carefully reconsidering the evidence, even when the change actually represents a framing-induced drop in accuracy. However, this does not mean that physician doubt was uniformly harmful. When the physician expressed doubt about an incorrect hypothesis, model accuracy increased modestly relative to the condition in which the same incorrect hypothesis was presented as the physician’s current impression. This suggests that in settings where a second medical opinion is sought, epistemic framing becomes a subtle safety issue. The model must challenge incorrect hypotheses while retaining correct ones when doubt is unjustified. In our exploratory analysis, we found that cases that were most fragile under neutral prompting were also the most sensitive to framing effects. This suggests that interactional risk may be concentrated precisely where decision support is most needed. Furthermore, a meaningful subset of unjustified responses was delivered with high confidence, particularly among unjustified-confirmation events, which could amplify their influence.

Our findings complement recent studies on the use of LLMs to assist physicians with reasoning and diagnostic tasks under structured workflow conditions (2,5,15). All three of the evaluated models demonstrate high performance on the MedQA task, achieving accuracy rates between 92% and 95% on the neutral baseline. This aligns with findings from other recent publications that have used this benchmark (16). However, our results highlight a vulnerability relevant to deployment: even when a model achieves high neutral accuracy, its output changes when the physician introduces a prior hypothesis or expresses doubt. This extends previous findings on workflow-sensitivity (5,20) by isolating a narrower mechanism: within a second-opinion interaction, the physician’s epistemic stance itself can alter the model’s accuracy. Importantly, our study also adds a complementary direction of influence to the literature on clinician–AI automation bias (12,13,14). Prior work examined how physicians are influenced by AI recommendations; we show that physician framing affects the AI response itself. In real clinical settings, both parties may act collaborate interactively, which independent accuracy evaluations of either entity do not capture.

Our results are consistent with evidence that conversational alignment can conflict with factual correctness. Ibrahim et al. showed that training models to be warmer reduces accuracy and amplifies sycophancy in medical question-answering (9), while Chen et al. demonstrated that LLMs may comply with inaccurate medical prompts when helpfulness overrides critical reasoning (17). However, our findings refine this literature in two ways. First, exact adoption of an incorrect physician hypothesis was less common in our setting (28 events), suggesting that frontier LLMs may resist direct confirmation of an erroneous clinical impression under structured second-opinion framing. Second, and more importantly, the dominant failure mode was rejection of a correct answer after physician doubt (86 events). This extends the concept of sycophancy beyond endorsement of incorrect beliefs: over-accommodation may also manifest as excessive deference to expert uncertainty. This is particularly important in the field of medicine as physician-framed prompts may carry authority in ambiguous cases when the model treats stated clinical hypotheses as meaningful diagnostic context (18).

These results also add to growing evidence that benchmark performance alone is insufficient for evaluating LLM reliability for medical tasks. Studies have shown that LLM performance in clinical decision-making is sensitive to instruction, information quantity, and information order (21). Furthermore, even strong standalone LLM performance does not translate to improved outcomes for general-public users (22). In expert settings with radiologists, workflow evidence and disagreement-prediction approaches also suggest that human–AI interaction shapes outcomes (5). When the same evidence yields different outputs depending solely on how the request was framed, our results also indicate that similar structural attention is needed for LLM integration in second opinion workflows. This finding is directly relevant to real-world practice, as there is evidence that physicians engage LLMs through diverse input approaches — generating, challenging, confirming, and refining hypotheses — meaning that confirmation-seeking and contradiction-seeking are not artificial prompt constructs but approximations of authentic querying behavior (8). The present findings align with other recent studies that call for independent, real-world evaluations of AI tools (16). While many publications focus solely on outcome and accuracy evaluations, we believe that the entire medical information flow should be investigated. This chain is shaped by interaction nuances on both the input and output sides, starting from possible variations in the patient input phase (23), and continuing across the processing phase, where physicians interact with the respective tools.

These findings have implications for the evaluation and deployment of LLMs in medical settings. For evaluation purposes, medical second opinions under variable physician-framed prompts are valuable scenarios for testing because they approximate how clinicians actually query these systems. Evaluations should distinguish between unjustified confirmation and unjustified rejection because these reflect different interactional vulnerabilities with real-world relevance. When deploying models and training physicians to use them, the influence of framing should also be considered. Interface designs that require the model to generate an independent case assessment before incorporating the physician’s hypothesis, or that explicitly separate supporting and contradicting evidence, may reduce inappropriate anchoring.

This study has several limitations. First, the cases were derived from MedQA-style multiple-choice vignettes rather than open-ended or multi-turn clinical scenarios. This format allowed for the controlled testing of causal epistemic framing while keeping the evidence constant. However, this simulation of second opinion scenarios does not fully replicate genuine clinical consultations, which are more ambiguous. Second, the physician framings were simulated and experimentally standardized. While this improves internal validity and enables a fully crossed study design, it does not capture the full variability of natural physician prompting. Third, the study evaluated three frontier models at a specific point in time. Although the models showed broadly similar directional patterns, the magnitude of framing sensitivity differed across models, and these findings may be transient and subject to change with model updates. Lastly, the study evaluated model outputs rather than the behavior of physicians after receiving the information. To increase the ecological validity of the results regarding unjustified confirmation or rejection, future studies should examine whether physicians recognize these framing-induced errors, and which factors mitigate their impact.

Overall, this study across three frontier LLMs shows that their accuracy in medical second-opinion settings is interaction-sensitive. Neutral benchmark accuracy did not sufficiently predict how the models would behave when presented with identical clinical evidence but different physician framings. The dominant safety-relevant failure mode was not the adoption of incorrect physician hypotheses but rather the rejection of correct answers when physicians expressed doubt. This finding broadens the scope of existing research on medical LLM sycophancy and clinician-AI collaboration by identifying an additional form of over-accommodation: deference to physician skepticism. Medical evaluations of LLMs should move beyond static benchmark accuracy and include real-world tests of the epistemic factors that clinicians may introduce.

## Materials and Methods

### Study design and preregistration

We conducted a preregistered, factorial experiment to evaluate whether physician epistemic framing affects the accuracy of LLMs in simulated medical second-opinion settings. The study was designed to evaluate two safety-relevant failure modes: unjustified confirmation, defined as adoption of an incorrect physician hypothesis, and unjustified rejection, defined as failure to retain the correct answer when the physician expresses doubt about it. The experiment used a fully crossed 2×2 within-case design. Each clinical case was evaluated under five prompting conditions by three LLMs. The primary unit of analysis was one model response for a given case, condition, and model. The study was preregistered prior to data generation on the Open Science Framework (OSF; available at https://osf.io/z72cf/overview).

### Clinical case dataset and preprocessing

The study used a preregistered sample of 500 clinical case vignettes in multiple-choice style derived from the MedQA dataset, following the sampling procedure and subset created by Ibrahim et al. (9). The preregistered input dataset contained 500 cases. For the final matched analysis, we retained the 499 cases for which responses were available from all three models across all five experimental conditions. Each item contained a clinical vignette, five answer options labelled A–E, and among those only one prespecified ground-truth correct answer. The source case format had previously been adapted from examination-style questions into conversational clinical queries by Ibrahim et al. Additionally, we aligned the vignettes with a physician-facing second-opinion scenario by replacing family-role terms such as “my brother/sister/mother/father” with “my patient”. No clinical cases were excluded after observing model performance unless a case was technically unusable due to absent answer options or processing errors.

### Experimental conditions

The experiment used a 2×2 factorial design with an additional neutral baseline condition. The two manipulated factors were the correctness of the physician’s stated hypothesis, defined as correct versus incorrect, and the physician’s epistemic framing, defined as confirmation-seeking versus contradiction-seeking. Condition-specific prompts were constructed as minimal linguistic contrasts to isolate the experimental manipulation while keeping all other wording as constant as possible to minimize prompt-related confounding. The neutral baseline condition did not include a physician hypothesis and served as the reference for estimating framing-related changes in model accuracy. Table 1 characterizes the five experimental conditions and their analytic role in testing unjustified confirmation and unjustified rejection.

**Table 1.** Experimental conditions for physician epistemic framing in medical second-opinion settings.

| Condition | Physician hypothesis | Epistemic framing | Prompt manipulation | Interpretation |
| --- | --- | --- | --- | --- |
| C0: Reference condition | None | Neutral | No physician hypothesis was provided; the model was asked to independently assess the case and identify the correct answer option. | Neutral baseline accuracy without physician framing |
| C1: Positive control | Correct | Confirmation-seeking | The physician’s stated impression was the correct answer option and was framed as the current clinical impression seeking<br>verification. | Beneficial confirmation of a correct physician hypothesis |
| C2:<br>Unjustified-confirmation | Incorrect | Confirmation-seeking | The physician's stated<br>impression was an<br>incorrect answer option<br>and was framed as the<br>current clinical<br>impression seeking<br>verification. | Adoption of an<br>incorrect<br>physician<br>hypothesis<br>under<br>confirmation-seeking framing |
| C3:<br>Unjustified-rejection | Correct | Contradiction-seeking | The physician initially<br>considered the correct<br>answer option but<br>expressed doubt about<br>its validity. | Failure to retain<br>the correct<br>answer under<br>physician<br>skepticism |
| C4:<br>Negative<br>control | Incorrect | Contradiction-seeking | The physician initially<br>considered an incorrect<br>answer option and<br>expressed doubt about<br>its validity. | Beneficial<br>correction when<br>physician<br>skepticism is<br>epistemically<br>appropriate |

### Model selection and prompt structure

We evaluated three state-of-the-art LLMs available via API from OpenAI [SDK version 2.15.0, model gpt-5.5, batch processing], Anthropic [SDK version 0.99.0, model claude-opus-4.7, batch processing] and Google [SDK version 1.75.0, model gemini-3.1-pro-preview]. All models were queried under comparable inference settings: temperature and “reasoning/thinking” settings were set to 0 (or to the closest equivalent), maximum output tokens were set to 1600. All models received an identical system prompt instructing them to act as a medical expert providing a second opinion to a physician, to select the correct answer from the given options, and to return a short explanation. Each query comprised the system prompt, a universal case block presenting the vignette and answer options A through E, and a condition-specific physician request instantiated with the relevant answer letter and text. The required response format was a structured JSON object with the final answer letter, a self-elicited confidence score between 0 to 100, and a concise physician facing explanation (for detailed settings see https://osf.io/z72cf/files/osfstorage).

### Outcome variables

The primary unit of analysis was a single model response for a given case, experimental condition, and model (case × condition × model), yielding a theoretical maximum of 7,500 model responses in total (500 cases × 5 conditions × 3 models). The final matched analytic dataset included 499 cases and 7,485 model responses. *Accuracy* was defined as a binary indicator equal to 1 when final_answer_letter matched the correct answer letter for a given case and 0 otherwise. *Unjustified confirmation* was defined as a binary indicator equal to 1 in C2 when final_answer_letter equaled the preregistered incorrect physician hypothesis, capturing exact adoption of an erroneous clinical impression. *Unjustified rejection* was defined as a binary indicator equal to 1 when a model answered a case correctly in the neutral baseline condition but failed to retain the correct answer in C3, where the physician expressed doubt about the correct hypothesis. *Beneficial confirmation* (C1) and *beneficial correction* (C4) were binary indicators capturing model accuracy in the respective control conditions. *Self-reported confidence* was treated as a descriptive variable and was not interpreted as a calibrated probability. An *overconfident unjustified response* was defined as an unjustified confirmation or unjustified rejection accompanied by a confidence rating of ≥80.

### Statistical analysis

Descriptive analyses reported the number of valid and invalid responses, accuracy, unjustified confirmation rate in C2, unjustified rejection rate in C3, beneficial confirmation rate in C1, beneficial correction rate in C4, mean and median self-reported confidence, and the proportion of overconfident unjustified responses. Results were summarized overall and stratified by model. Proportions were reported with 95% confidence intervals. The primary inferential analyses used logistic regression with cluster-robust standard errors at the case level to account for repeated observations of the same clinical cases across conditions and models. For accuracy, the dependent variable was binary correctness. The primary model included condition and model as fixed effects. A condition-by-model interaction model was fitted to examine model-specific framing susceptibility. The neutral baseline condition C0 served as the reference category. The two co-primary contrasts were C2 versus C0 for the unjustified-confirmation effect and C3 versus C0 for the unjustified-rejection effect. We report odds ratios, 95% confidence intervals, and p-values. To exploit the paired within-case structure, robustness checks were conducted separately for each model using McNemar tests for C0 versus C2 and C0 versus C3. We also summarized transition patterns, including cases changing from correct in C0 to incorrect in the framed condition and from incorrect in C0 to correct in the framed condition. Secondary analyses compared accuracy in C4 with accuracy in C2 to describe beneficial skepticism when the physician doubted an incorrect hypothesis. Model differences in framing susceptibility were evaluated using the condition-by-model interaction and by reporting model-specific accuracy differences, transition counts, and unjustified response rates. Case difficulty was operationalized exploratorily from C0 performance aggregated across all three models: cases answered correctly by all three models in C0 were classified as *clear*; cases for which at least one model answered incorrectly in C0 were classified as *ambiguous*. Two implementation decisions deviated from the preregistered analysis plan: we used logistic regression with cluster-robust standard errors at the case level instead of mixed-effects logistic regression, and we operationalized unjustified rejection as the stricter paired transition from a correct answer in C0 to an incorrect answer in C3, while the broader C3 incorrect rate was retained in the accuracy analysis. All analyses were conducted in Python 3.13.

## Data Availability

All prompts, inference settings, analysis code, and datasets are publicly available on the OSF (https://osf.io/z72cf/files/osfstorage).

## Ethics

This preregistered study analyzed LLM-generated output based on public, benchmark-derived clinical vignettes. This experimental setup did therefore not involve any human participants or identifiable personal data.

## Author Contributions

FR: Conceptualization, Methodology, Software, Formal Analysis, Investigation, Data Curation, Writing (Original Draft, Review & Editing), Visualization, Project Administration; WK: Conceptualization, Methodology, Field Expertise in Human-AI Interaction, Writing (Original Draft, Review & Editing), Supervision; FB: Conceptualization, Field Expertise in Physician-AI Interaction, Resources, Writing (Original Draft, Review & Editing), Supervision; SDB: Conceptualization, Software, Validation, Investigation, Resources, Data Curation Provision, Writing (Original Draft, Review & Editing), Supervision, Project Administration

